# Lead exposure from general aviation emissions in the UK: a review and call for action

**DOI:** 10.1101/2022.07.12.22277256

**Authors:** Ashley Mills, Stephen Peckham

**Affiliations:** Centre for Health Services Studies, The University of Kent, George Allen Wing, Cornwallis Building, Canterbury, Kent, CT2 7NF, UK

**Keywords:** lead poisoning, aviation, leaded fuel, air quality

## Abstract

Leaded fuel emissions exposed a global population of children to lead and its profound health consequences. Recognition of its harms precipitated a global phase out and replacement with unleaded substitutes for road vehicles. Despite this widespread recognition and action, aviation fuel for piston engine aircraft still contains lead. Leaded aviation fuel (AVGAS100LL) contains 0.56g of tetraethyl lead per litre and this lead must be jettisoned from the engine during operation to prevent fouling. This action distributes lead and lead compounds into the air and soil around general aviation airports. This has been shown to increase the blood lead levels of children living nearby to clinically significant levels. Whilst this problem is recognised by the EPA in the U.S, it has received little attention in the UK. We provide a review of the situation in the UK with regard to the current policy and regulation framework. We analysed the UK’s general aviation airport fleet, general avation airport data, and GB residential address data. We estimate the unleaded-readiness of the UK aviation fleet and the current usage of fuels at UK general aviation airports. We provide a first order estimate of the number of residential addresses exposed to lead near general aviation airports. We find that the majority of aviation fuel sold in the UK is leaded and that there are 370721 residences within 4km of a general aviation airport at risk from exposure to lead emissions. Finally we present a path forward for regulation change and public health monitoring.

## 1 Introduction

Lead is a toxic heavy metal that has no role in human physiology. Natural human levels of lead should be very close to zero [1]. Lead is toxic to all, but children are particularly vulnerable for a variety of factors: children absorb 4-5 times more environmental lead than adults [2], have undeveloped renal physiology, exhibit higher oral exploratory behaviour, have an undeveloped blood/brain barrier, are subjects of extremely sensitive developmental processes, and being young have a longer period of time for effects to be felt [3]. Lead is stored in the skeletal system and is a cumulative toxin causing irreversible and untreatable damage.

The toxic effects on children are marked to occur at extremely low concentrations and there is believed to be no threshold for harmful effects [3, 4, 5]. The relationship between lead and IQ appears to be non-linear with initial increments in blood lead levels (BLLs) having a higher impact than later increments: a study looking at BLLs of children over a period of 5 years and IQ measured at 3 and 5 years of age [6] estimated a 7.4 point drop in IQ as BLLs increased from 0-10 µg/dL and a further 2.5 point drop between 10 and 30 µg/dL. It is believed that as levels increase cell protective mechanisms activate which attenuate the incremental damage. It is imperative therefore that children are not exposed to lead in any quantity.

Lead exposure comes from a variety of sources [4]: industrial emissions; leaded paint; lead pipes; leaded toys and equipment; ceramic glazes; e-wastes; traditional medicines; contaminated soil; as well as leaded fuel.

Leaded petrol has caused more human exposure to lead than any other source worldwide [7]. As an illustration of the magnitude of its impact: abatement of lead in petrol in the US is believed to have increased the average intelligence of American children by between 2-5 IQ points since 1980 and yielded an aggregate economic benefit of over $6 trillion (as of 2017) [1].

After a 20 year campaign to remove lead from petrol, in 2021, the United Nations Environment Programme (UNEP) Partnership for Clean Fuels and Vehicles announced that leaded petrol has been removed from the supply chain in every country of the world for the first time since before its introduction in 1923 [8]. It is noteworthy however that current legislation for “unleaded” petrol still permits upto 5mg of lead per litre [9], which could amount globally to hundreds of tons of emitted lead per annum at current consumption rates.

Despite this success, and despite universal acceptance of the toxicity of lead, the most common aviation fuel used worldwide for general aviation (AVGAS100LL) still contains high quantities of lead.

### 1.1 Leaded aviation fuel (AVGAS)

Lead is added to aviation fuel (AVGAS) in the form of tetraethyl lead (TEL) to satisfy the requirements of Piston Engine Aircraft. TEL prevents engine knocking (early detonation), lubricates engine components, and protects recession of intake and exhaust valves [10]

During combustion TEL is converted to lead oxide. This must be catalysed into brominated lead oxybromides by another additive called Ethylene Dibromide (EDB), to prevent the accumulation of lead oxide on engine parts (engine fouling). EDB is a pesticide.

At high levels of exposure, EDB is lethally toxic to a broad variety of insects and animals including humans [11] and chronic occupational exposure of EDB has been shown to affect fertility [12]. EDB can persist for decades in soil and in aquifers [13], and yet little is known about the health effects of long-term low-level exposure. In 2019 the EPA designated EDB a *“high priority substance for risk evaluation”* and is in the process of preparing this evaluation [14].

The catalysed lead oxybromides are exhausted from the engine and enter the atmosphere where they condense as fine particulates. These particulates distribute around a mean of 1µ*m* in diameter and particle concentrations are 3x higher for take-off and 2x higher for climb flight phases relative to cruising [15]. Thus more lead is exhausted nearer to airports per unit of time than further away from them.

In response to health concerns, lead in AVGAS has been successively reduced and a so-called *“low lead”* variant (AVGAS100LL) is now widely used. Despite being called *“low lead”* AVGAS100LL contains 0.56g of TEL per litre [16].

### 1.2 The push for unleaded AVGAS

In the US, significant campaigns have been mounted by environmental groups to petition against leaded AVGAS use. In 2011 the Center for Environmental Health (CEH) sued a number of AVGAS suppliers and producers under the *“Safe Drinking Water and Toxic Enforcement Act”* for failing to warn residents near airports in California about exposure to lead. CEH won the lawsuit in 2014 with a settlement [17] that resulted in 24 airports being required to notify all residents within 1km of the hazard, and to erect warning signs at each airport with the following text:

#### WARNING

> *The area within one kilometer of this airport contains lead, a chemical known to the State of California to cause cancer, birth defects or other reproductive harm. Lead is contained in the aviation fuel (“Avgas”) that is used by small piston engine aircraft that take off and land at this airport. People living, working, or travelling near this location will be exposed to lead as aircraft take off and land*.
>
> *For more information, visit www.ceh.org/avgas* “

The environmental group Friends of the Earth sued the EPA in 2012, leading them to issue a declaration to make a final ruling on an “endangerment finding” for leaded AVGAS by 2018 [18]. At the time of publishing they had not issued a ruling.

Since 2011, the Federal Aviation Association (FAA) in the US has been running a program called the Piston Aviation Fuels Initiative (PAFI) whose goal is to help develop and test unleaded aviation fuels to replace AVGAS100LL [19]. As of writing, this is an ongoing project that has not delivered a suitable replacement for AVGAS100LL that works with all of the operating fleet.

Note that an unleaded AVGAS called UL91 exists, but as the name implies, has a lower octane rating. UL91 is not suitable for all existing engines, and has neither been widely adopted nor is widely available (see below section on AVGAS in the UK).

Outside of the PAFI process, independent research has been carried out. It was announced in July 2021 that a 100 octane unleaded fuel had been developed by General Aviation Modifications Inc (GAMI) and received certification by the FAA [20]. GAMI claims that the fuel, known as G100UL, will provide a *“functional fleet-wide drop-in replacement for 100LL”* although admitting the fuel will cost more than 100LL. It remains to be seen how quickly certification can be expanded to a wide variety of engine types and whether adoption will be widespread.

### 1.3 AVGAS, airports, and child blood lead levels

There are to our knowledge only two studies examining child blood lead levels and their association with airport proximity. In a groundbreaking study [21], the first of its kind, researchers examined child blood lead levels (BLLs) of 125,197 children living in geospatial proximity to airports using leaded AVGAS in North Carolina. They found significant increases in BLLs for children living within 1.5km of an airport, with an inverse dose-response observed with distance.

A more ambitious study was conducted on a 2015 dataset from Michigan that looked at the BLLs of over 1 million children [22] in proximity to airports with between 84 and 13188 movements per year. Child BLLs were elevated close to airports and a dose-response effect was observed with regard to airport proximity as well as airport traffic volume. The effects were statistically significant out to 3km away from the airport but a non-statistically significant effect was observable out to 4km. The study found that being downwind of an airport amplified exposure as did the number of takeoffs/landings at a given airport.

Children were 25.2% more likely to have a BLL in excess of 5 µg/dL within 1km of a piston engine airport relative to a reference considered to be *>* 4km away. They were 16.5% more likely at a distance between 1-2km, 9.1% more likely at a distance of between 2-3km, and 5.4% more likely at a distance of 3-4km. Beyond this the impact of the airport was found to level off.

The exposure pathway in this case twofold: direct inhalation as a result of ongoing deposition from aviation traffic, and inhalation of lead contaimnated dusts from re-suspension of previous soil deposition. It is likely that there is direct indoor deposition through air ingress as is the case with traffic and industrial lead sources [23], and dusts will be tracked indoors contributing to indoor exposure.

### 1.4 Leaded AVGAS in the UK

The Department For Transport (DfT) publishes annual fuel consumption data for different grades of fuel including “Aviation spirit” (AVGAS) [24]. Unfortunately the DfT does not differentiate between leaded and unleaded AVGAS grades. However, as will be seen below, the majority of AVGAS used in the UK is leaded so some idea of the volumes involved can be understood from the data.

The data are published in millions of tonnes. The density of AVGAS varies depending on refining practices, storage temperature, and other factors, but is typically stated in material data safety sheets as being between 0.68 and 0.74 kg/litre [25, 26]. We have assumed the midpoint of 0.71 kg/litre density as a representative value, and a value of 0.56g/litre for lead content [22]. We have assumed it all to be leaded as an upper bound of exposure.

Figure 1 plots the annual AVGAS consumption for the period 1998 to 2019 (data are not available before 1998) along with the associated maximum cumulative lead emissions.

**Fig. 1.**
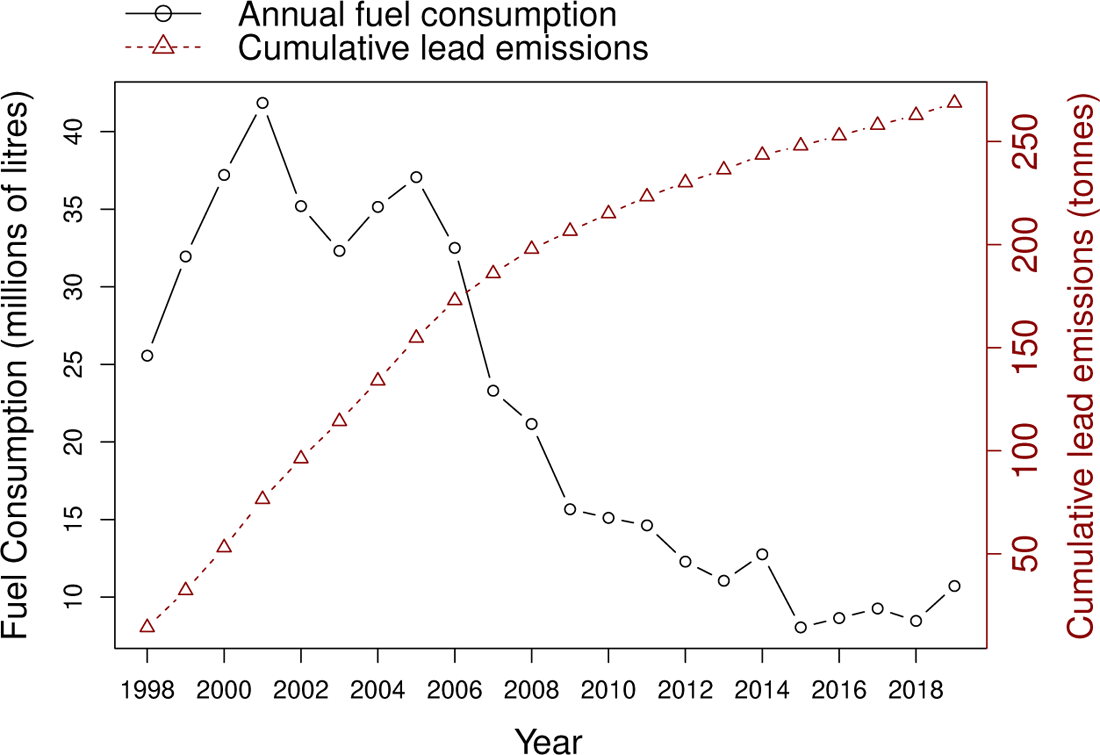
Annual AVGAS consumption in the UK for the period 1998-2019 against estimated maximum cumulative lead emissions.

As can be seen, AVGAS consumption has broadly declined over the last 20 years, and although a recent upswing has been observed it must be noted that the DfT figure for 2019 is stated as being an estimate. Cumulative estimated maximum lead emissions amount to over 250 tonnes with a mean emission of just over 5 tonnes per annum in the last 5 years. Actual figures depend on how much of the AVGAS used is leaded VS unleaded, and the amount actually used in planes VS that discarded as waste. We could find no way to estimate how much AVGAS is discarded with publicly available data.

Since neither the DfT or HMRC distinguish between the unleaded and leaded grades of AVGAS we examined availability of leaded and unleaded AVGAS formulations at UK aerodromes to understand the likely proportional usage.

A list of 124 currently licensed and certified (as of June 2021) aerodromes were obtained by contacting the UK’s aviation regulator: the Civil Aviation Authority (CAA) [27]. Data for each aerodrome was obtained by web-scraping the Aeronautical Information Service (NATS) eAIS package [28] which has an effective publication date of 14/07/21. The locations and licence type of each of these aerodromes are shown in Figure 2.

**Fig. 2.**
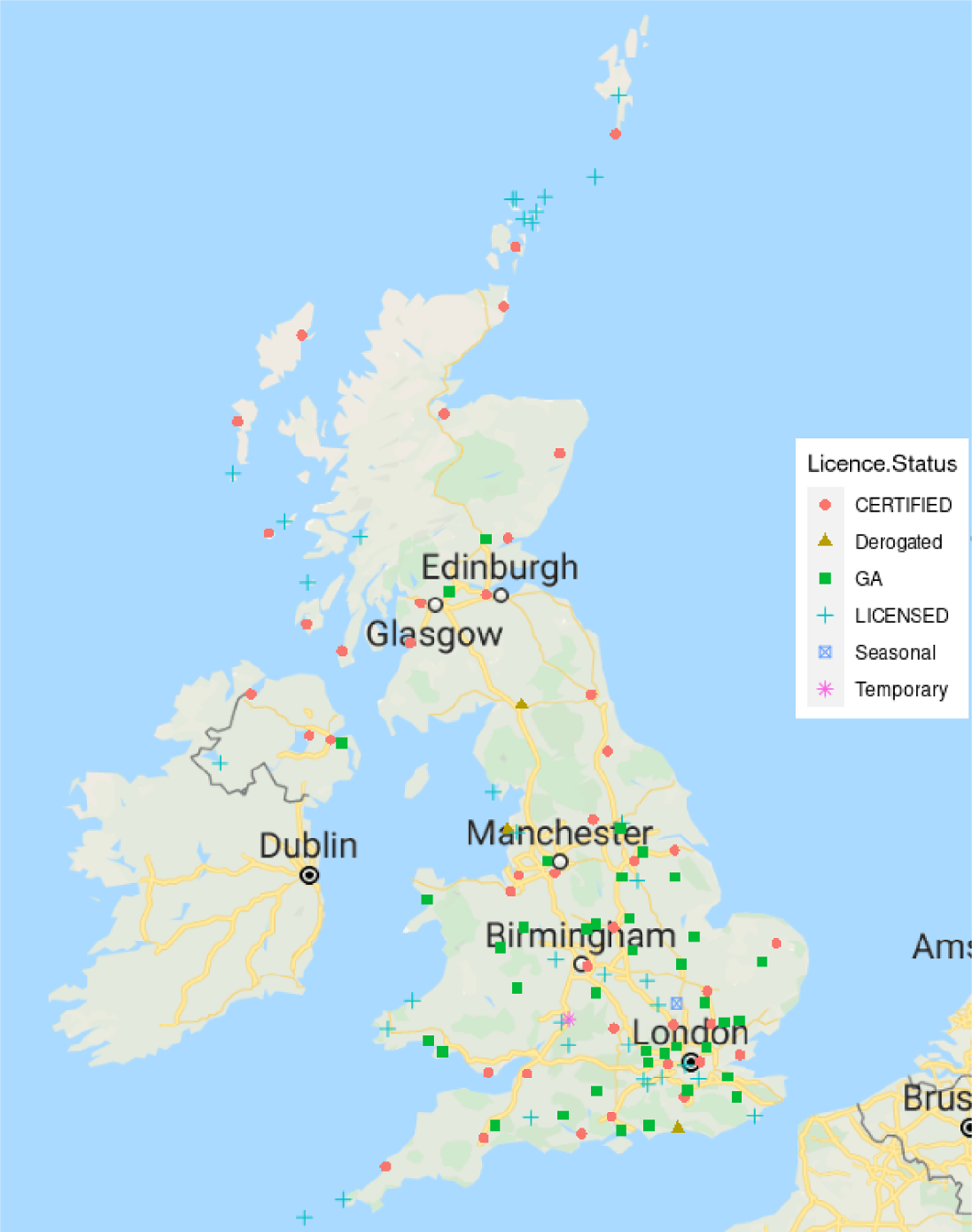
Locations and licence types of CAA regulated aerodromes in the UK

For each airport we extracted the “Fuel and oil types” and “Fuelling facilities/capacity” fields from the “HANDLING SERVICES AND FACILITIES” section of each entry. Leaded AVGAS availability was determined by detecting the string “AVGAS 100”. Unleaded AVGAS availability was detected by detecting the strings “UL91” and “UL-91”. Detections were confirmed manually via a generated summary table containing the extracted fields.

A total of 107 aerodromes had non-null “Fuel and oil types” entries, and 62 aerodromes had non-null “Fuelling facilities/capacity” entries. Table 1 summarises this data by aerodrome licence type in terms of the number of aerodromes in each category providing 100LL and UL91 fuels.

**Table 1.**
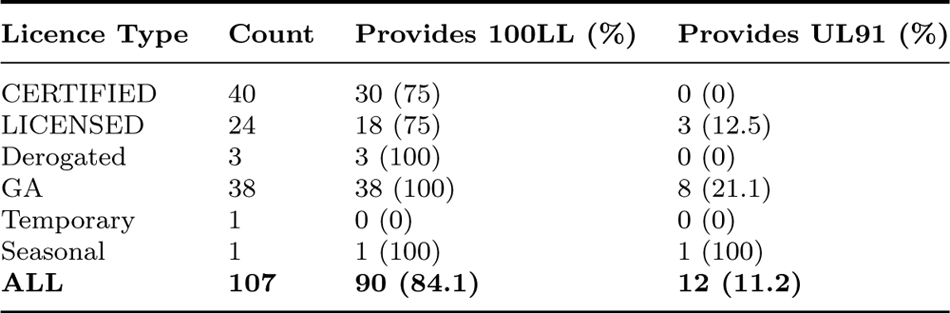
Number of aerodromes providing 100LL and UL91 fuels stratified by aerodrome licence type.

| Licence Type | Count | Provides 100LL (%) | Provides UL91 (%) |
| --- | --- | --- | --- |
| CERTIFIED | 40 | 30 (75) | 0 (0) |
| LICENSED | 24 | 18 (75) | 3 (12.5) |
| Derogated | 3 | 3 (100) | 0 (0) |
| GA | 38 | 38 (100) | 8 (21.1) |
| Temporary | 1 | 0 (0) | 0 (0) |
| Seasonal | 1 | 1 (100) | 1 (100) |
| **ALL** | **107** | **90 (84.1)** | **12 (11.2)** |

Licensed airports are those which permit public transport and commercial passenger flights, and those which are certified permit larger numbers of passengers and runway lengths. Certified airports tend to be the large national public airports. Derogated airports are those which should be certified because they meet other necessary conditions but due to low passenger numbers have been given a waiver. GA airports are all other non-commercial aerodromes, typically used for flight training, parachute jumping, and leisure flights. Temporary licenses are issued to airports for specific public access: for example Cheltenham Heliport (EGBC) is temporarily licensed for the Cheltenham Festival each year. A seasonal airport is licensed only during nominated days: for example Old Warden (EGTH) has a license for only 20 days per year which it uses primarilty for airshow days.

Overall, 84.1% of the 107 aerodromes who specify this information, provided 100LL and only 11.2% provided UL91. All the GA aerodromes provided 100LL and 21.1% of them provided UL91. We don’t know what the fuel provisions are for the remaining 17 airports, but it should be clear that even if none of them provided 100LL and all of them provided UL91, the vast majority of fuel available would still be 100LL.

Only 21 aerodromes contain information about fuel capacities regarding 100LL and UL91. Nine of these state capacity for a generic “AVGAS”, but only one of these provides UL91 (Gloucestershire) so the rest are assumed to be 100LL. Table 2 summarises the available capacity information for the 20 aerodromes that provide unambiguous data.

**Table 2.**
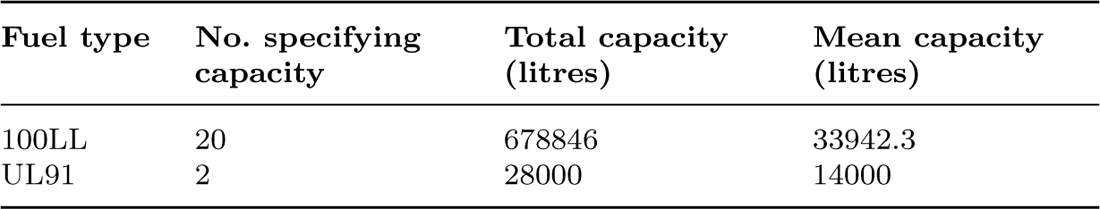
Fuel capacity for 100LL and UL91 for UK aerodrome that provide this information (20 aerodromes)

| Fuel type | No. specifying capacity | Total capacity (litres) | Mean capacity (litres) |
| --- | --- | --- | --- |
| 100LL | 20 | 678846 | 33942.3 |
| UL91 | 2 | 28000 | 14000 |

Far more capacity is specified for 100LL relative to UL91, and the mean capacity is higher for 100LL than UL91. In the two aerodromes (EGLK and EGHA) where both capacities are specified, 100LL capacity is 3.4 and 1.4 times greater than UL91 respectively.

From this evidence, it is reasonable to conclude that the vast-majority of AVGAS used is 100LL.

### 1.5 UK fleet fuel capabilities

A list of registered and airworthy piston-engine fixed-wing light aircraft was obtained from the CAA (direct contact). A fixed-wing light aircraft has a maximum take off weight of less than 5650kg.

The dataset contained a total of 5487 piston-engined aircraft. We identified 640 different engine types from 91 manufacturers, and for each engine type an attempt was made to manually identify the permitted fuel types.

The top three engine manufacturers were Lycoming (2679 aircraft), Continental Motors (893 aircraft), and Rotax (711 aircraft), accounting for 78% of the fleet. Fortunately, these manufacturers provided service bulletins which identified permitted fuels for the majority of their engine variants [29, 30, 31]. Permitted fuels for other engines were manually sought out, either from manufacturer websites, or where the European Union Aviation Safety Authority (EASA) provided a Type Certification document [32]. Failing this, Wikipedia, or flying forum materials were consulted.

We were able to identify the permitted fuel for a total of 4817 aircraft (87% of the fleet). Information was unavailable for 676 aircraft. Table 3 summarises the results of the analysis.

**Table 3.** Permitted fuels by aircraft number in the UK aviation fleet

| Fuel Type Permitted? | Number of aircraft (%) |
| --- | --- |
| Some form of unleaded fuel | 3271 (59%) |
| 100LL | 4626 (84%) |
| Other (high lead, legacy) | 184 (3%) |
| Information unobtained | 670 (12%) |

Note that the second column doesn’t total to 100% because aircraft are certified to use multiple fuels. 59% of aircraft were identified as being able to use some form of unleaded fuel (UL91 or a high octane unleaded petrol), and 28% were identified as not being able to. This is inline with general statements made by BP in its UL91 fuel brochure [33] that *“UL91 is suitable for about 55% of planes”*. 84% of aircraft could use 100LL, and 3% could not.

Information for 12% of the fleet was missing, but it is clear that the majority of the fleet can use unleaded fuel. The issue seems to be unavailability of this fuel, and a lack of demand. Notwithstanding this, there is still a sizable portion of the fleet unable to use leaded fuel.

### 1.6 Exposure risk for leaded fuel emissions for GB residential addresses

The work of [21] and [22] demonstrated statistically significantly affects on child BLLS of airport lead emissions for children living within 3km of an aerodrome, and an observable affect upto 4km.

Taking 4km as a maximum reasonable distance for “at risk”, as an average over meteorological and runway parameters, we sought to estimate the number of residences “at risk” of exposure to lead from UK aviation (or in this case GB as explained below).

GB (UK excluding Northern Ireland) residential address data was obtained from Ordnance Survey via a research license for the AddressBase Core product [34]. This contains 33,596,527 addresses with centroids, labelled according to address category (residential, business etc) as well as various other data. Northern Ireland data was not available from Ordnance Survey.

The centroids of the main runway for each GB airport was obtained from NATS eAIS [28] as above.

For each airport centroid, circular polygons were generated to establish rings at 1km, 2km, 3km and 4km.

The number of residential properties in each ring can be established by computing the intersection of the address data and the rings. It is however computationally inefficient to perform this directly, given the large number of addresses. Therefore, postcode sector polygons were obtained from The University of Edinburgh’s datashare platform [35] in order to enable a hierarchical computational approach. The mechanism followed is outlined in Algorithm 1.

In practice this only needs to be executed for the 4km ring, as the smaller rings are subsets and can be easily computed from the former data.

As an example, Figure 3 shows the locations of residential addresses around Headcorn/Lashenden (EGKH) airport within 1km, 2km, 3km, and 4km rings around the main runway centroid.

**Fig. 3.**
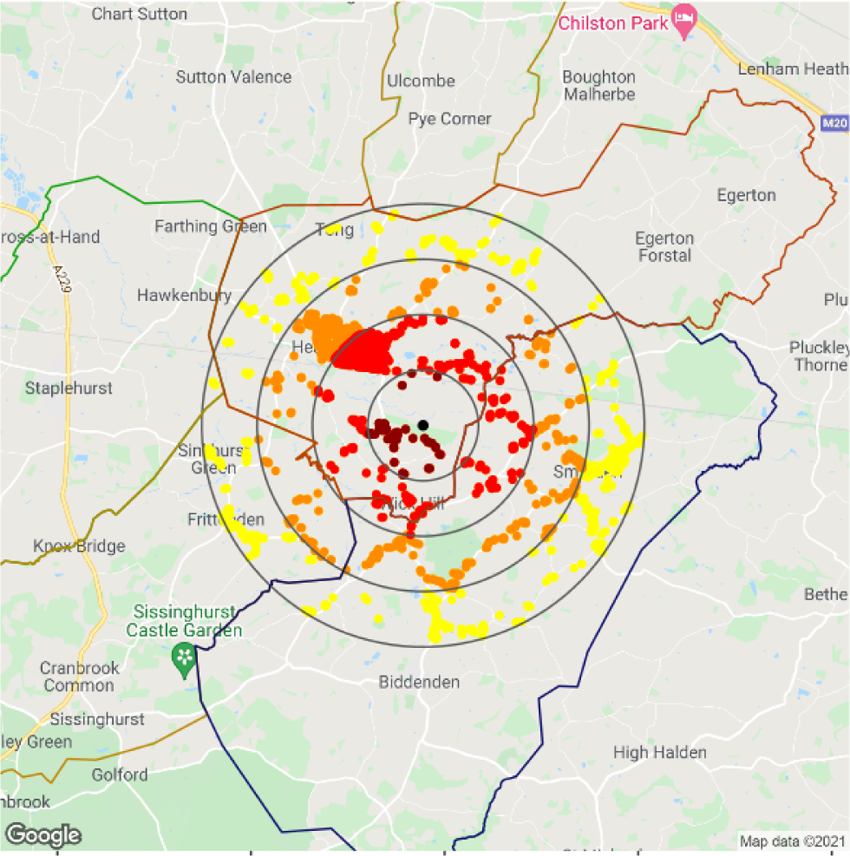
1km, 2km, 3km and 4km rings around Headcorn/Lashenden aerodrome (EGKH) with residential addresses plotted as coloured points. Different colors are used for each ring. Postcode sector boundaries are also shown. Point data © Crown copyright and database right 2021. Postcode sector data © see data acknowledgement section. Background map data © Google LLC 2021.

#### Algorithm 1 An algorithm to extract residential addresses “at risk” of lead exposure from GB aerodromes

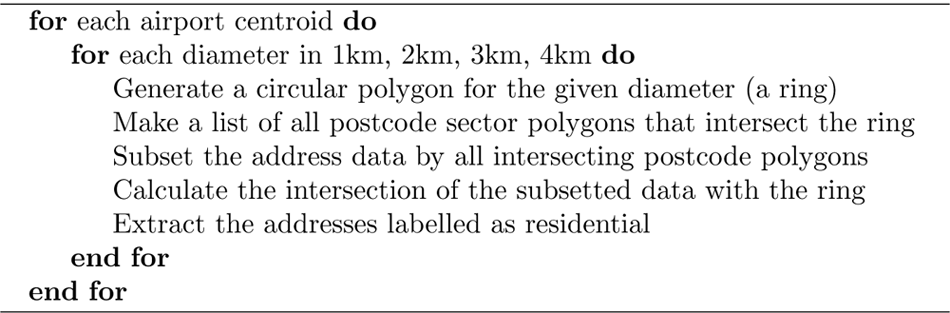

Table 4 shows the mean number of residential households within different sized ring radii, as averaged over the dataset of all airports, for each airport type.

**Table 4.** Mean number of residences for each ring diameter and airport type, rounded to nearest whole number.

| Airport Type | 1km | 2km | 3km | 4km |
| --- | --- | --- | --- | --- |
| CERTIFIED | 374 | 3378 | 9897 | 20484 |
| LICENSED | 1190 | 4631 | 10470 | 19339 |
| Derogated | 681 | 8209 | 16871 | 24592 |
| GA | 265 | 1749 | 5251 | 10019 |
| Temporary | 268 | 6018 | 23082 | 37333 |
| Seasonal | 53 | 561 | 1448 | 4485 |
| **ALL** | **547** | **3265** | **8675** | **16699** |

Table 5 shows the sum of residential households within different sized ring radii, as summed over the dataset of all airports, for each airport type.

**Table 5.** Sum of residences within different sized rings around airports, for different airport types.

| Airport Type | 1km | 2km | 3km | 4km |
| --- | --- | --- | --- | --- |
| CERTIFIED | 14200 | 128380 | 376101 | 778373 |
| LICENSED | 32120 | 125041 | 282695 | 522158 |
| Derogated | 2043 | 24626 | 50613 | 73775 |
| GA | 9796 | 64714 | 194273 | 370721 |
| Temporary | 268 | 6018 | 23082 | 37333 |
| Seasonal | 53 | 561 | 1448 | 4485 |
| **ALL** | **58480** | **349340** | **928212** | **1786845** |

From Table 5 we can see that for 194,273 residences near to GA airports, children living in these houses are likely to have elevated blood lead levels.

Approx 370,721 residences are at risk of being exposed to damaging levels of lead, just from GA airports where the majority of traffic is piston prop aircraft.

## 2 Discussion

The purpose of this paper is to highlight the issue of aviation lead emissions in the UK which has received little or no attention. We have provided a first-order estimate of the number of residences at risk of lead exposure, and analysed the UK GA fleet and fueling facilities. There are a number of limitations with this work that need discussion.

Firstly, our metric for “at risk” is based on a simple distance metric from the centroid of each airport runway. This is based on findings from two US studies that looked at child BLLs and airport proximity, and in particular the Michigan study [22] that looked at over 1 million children. The categorical BLL elevation probabilities provided therein are an average over the 448 airports they examined, and the average monthly movements of these airports was 406.09. The same study also found that children residing downwind of an airport had a higher probability of elevated BLL, as did living near an airport with a greater number of movements.

By applying a simple judgement such as “any residence within 4km is at risk”, we ignore the differences between airports in terms of movement numbers, and the location of residences relative to prevailing wind directions. It is almost certainly the case that many airports in the UK have more than 406.09 movements per month, including many GA airports and thus in many instances the risk will be underestimated. Likewise, there will be instances where the risk is overestimated.

Without accurate figures for number of flight movements for a given airport it isn’t possible to perform any kind of scaling for this. Small aircraft pilots are not required to register their flights as they can operate on Visual Flight Rules (VFR) only, and do not require a transponder, thus there is no official record for these flights. GA airports are not required to report how many movements per year occur at their facilities.

The residence address data we used does not specify which residences contain children, so the specific risk to children is unknown, and yet this is the most vulnerable category concerning exposure. We can only say that a residential address may contain children. It would be useful for the UK government to undertake a survey of residences near to aerodromes to rectify this information deficit. Furthermore we do not have information about soil lead concentrations in the UK around aviation sites, and no data concerning child BLL levels near to aviation sites are available.

These shortcomings highlight the lack of oversight of GA facilities and their impact on the health of local populations.

Finally, we have presented data for a wide variety of airport types, but have presented our conclusions in terms of GA airports, since it is at GA airports that piston engine aircraft, and thus lead emissions, abound.

The military is a user of leaded aviation fuel, and military aerodromes are not GA aerodromes, thus when speaking of risk from GA aerodromes we neglect the risk from military aerodromes. Furthermore, it also is not clear what impact the other types of airport have on health, where a mix of fuels is used, and where there are different restrictions on the types of aircraft allowed. For example, many of these airports permit helicopters, and many helicopters are piston powered.

Thus, whilst we can say for certain, that the GA airports pose a risk to health, the risk posed by all the other airport types remains unquantified. This risk should not be neglected and future work needs to be done to understand it.

In general the UK government recognises the importance of quantifying annual lead emissions and maintains the National Atmospheric Emissions Inventory (NAEI) for this purpose and to track other forms of air pollution [36]. In the course of this research however we examined their data and discovered they were using an emissions factor for lead of 0.0003924 gram/litre for aviation spirit. This is 1427 times less than the general figure accepted for lead in aviation spirit of 0.56 gram/litre. Upon contacting the NAEI about this they admitted the mistake and are in the process of updating the relevant publications and derived figures. This demonstrates the lack of scrutiny that has been applied to the issue of lead in aviation fuel in the UK.

### 2.0.1 The switch to unleaded aviation fuel

The UK aviation fleet needs to transition to unleaded fuel, and policy needs to be enacted to enforce this. We have shown that the majority of the existing fleet can use UL91, yet the majority of UK aerodromes do not stock this fuel. In the absence of availability of a 100UL fuel, legislation should be promoted to ensure that aerodromes stock UL91 and its use should be promoted over 100LL through government subsidy.

At the present time HMRC does not distinguish between leaded and unleaded fuels when tracking duty, which is a neglectful oversight given the health implications of leaded fuel. A clear method to proceed would be for HMRC to track aviation fuel by fuel type and differentiate the duty paid. Leaded fuels should have increased duty to dis-incentivise use. By tracking duty by fuel grade, the transition to unleaded fuel can then be quantized and judged against targets.

If G100UL can be shown to live upto its claims, then any aircraft that currently uses 100LL will be able to switch to an unleaded alternative. GAMI claims that G100UL, whilst likely costing 60-80 cents more than 100LL, will reduce engine maintenance intervals due to the lack of lead-fouling [37]. GAMI expects the rollout and certification process to take several years.

Once a 100UL fuel is available, and the additives have been shown to be safer than lead, legislation should be enacted to phase out 100LL and enforce adoption of the 100UL. Legislation is likely to be required since the additional cost of G100UL may discourage voluntary switching. Again, duty could be used as a mechanism to incentivise change.

### 2.0.2 Monitoring of child blood lead levels in the UK

There is no active surveillance of child BLLs in the UK. A passive laboratory-based monitoring program was piloted in 2014 across the UK in association with the British Paediatric Surveillance Unit (BPSU) [3, 38] and made permanent for England only in 2016.

It is now known as the Lead Exposure in Children Surveillance System (LEICSS) and the UK government produces annual surveillance reports [39]. LEICCS uses two passive, and voluntary, sources of information:

1. When a UK Accreditation Service (UKAS) accredited biochemistry or toxicology laboratory measures (and voluntarily reports) a BLL of an English resident child (defined as under 16 years of age) having a value ≥ 10 µg/dL.
2. Periodic searches of HPZone by the LEICCS organisation for such defined cases that were not reported by a laboratory. HPZone is a case management tool used by public health protection teams in England, and cases are voluntarily entered into this by users.

The original pilot study utilised laboratories and public health organisations in England, Scotland, Wales, Northern Ireland, and the Republic of Ireland. LEICCS is focused on Public Health England and English residents.

The original pilot study for LEICCS employed a third data source: BPSU registered paediatricians in all regions were asked to report monthly via a card system and identified cases [38]

Note that the case definition employed by LEICCS: a blood lead threshold of 10 µg/dL, is not aligned with contemporary understanding of harms which posit no lower effect limit. Toward this position, a recent report by PHE [40] for LEICSS recommends lowering of the case definition threshold for children to 5 µg/dL.

Passive sampling by nature, ad-hoc and incomplete. We have demonstrated that there are significant numbers of residential households at risk from lead exposure from aerodromes the UK. It should be considered a public health imperative to determine the nature and extent of this risk quantitatively, by measuring BLLs.

We propose a comprehensive active sampling program around UK aerodromes so that their risk can be properly understood in a public health context.

## 3 Conclusion

Lead emissions from aviation *is* a problem which affects the UK. Aerodromes are located sufficiently close to residential areas to pose a risk to children in those residences. The majority of fuel used by GA aircraft is leaded and yet the majority of registered piston engine aircraft (59%) are capable of using unleaded fuel in some form. A sizeable portion (41%) however remain unable to use all currently available unleaded fuels. A new unleaded high-octane (G100UL) fuel has recently become available that may serve to allow for a complete transition to unleaded fuels. It is likely that a duty incentive, or new regulation, would be required to drive this change.

The exposure of children to lead is acknowledged in the UK as being an ongoing problem, and yet is only measured via a passive monitoring system. There is no active monitoring of blood lead levels around known sources of emitted lead or indeed aerodromes. It is important for public health that the actual exposure of children to lead from aviation is understood and that an active monitoring system is established to achieve this goal.

This publication and its analysis, incomplete by virtue of lack of measured data in the UK, should raise awareness of the seriousness of the last remaining vehicular source of lead emissions in the UK. The UK authorities need to establish proper accounting for leaded fuels, take action toward their final phase-out, and implement mechanisms to monitor public health with regard to their impact.

## Data Availability

The postcode sector polygon data are available from the University of Edinburgh DataShare platform (https://doi.org/10.7488/ds/1947). This is a publicly available dataset.
The residential address data used were obtained by license from the Ordnance Survey via their AddressBase Core product (https://www.ordnancesurvey.co.uk/business-government/products/addressbase-core). This is a paid-for product but is available under a free license for research purposes, on a per-application basis.
The list of registered UK aircraft and their details can be obtained by contacting the CAA directly (https://www.caa.co.uk/home/).

https://www.ordnancesurvey.co.uk/business-government/products/addressbase-core

## Declarations

### Author contributions

All authors contributed to the study conception and design. Material preparation, data collection and analysis were performed by Ashley Mills. The first draft of the manuscript was written by Ashley Mills and all authors commented on previous versions of the manuscript. All authors read and approved the final manuscript.

### Ethics approval

Ethics approval was not required for this desk-based study.

### Consent to participate

Not applicable

### Consent for publication

Not applicable

### Availability of data and materials

The postcode sector polygon data are available from the University of Edinburgh DataShare platform [35]. These data are publicly available. The data were used here under provision of inclusion of the following licensing text:

> Postal Boundaries © GeoLytix copyright and database right 2012 Contains Ordnance Survey data © Crown copyright and database right 2012 Contains Royal Mail data © Royal Mail copyright and database right 2012 Contains National Statistics data © Crown copyright and database right 2012. GIS vector data. This dataset was first accessioned in the EDINA ShareGeo Open repository on 2014-03-14 and migrated to Edinburgh DataShare on 2017-02-22

The residential address data used were obtained by license from the Ordnance Survey via their AddressBase Core product [34] and was used under provision of inclusion of the following licensing text:

> © Crown copyright and/or database right 2021 OS. © Local Government Information House Limited Copyright and database rights 2021 100034829. This data contains data created and maintained by the Scottish Local Government.

The AddressBase Core dataset is a paid-for product but is available under a free license for research purposes, on a per-application basis.

The list of registered UK aircraft and their details can be obtained by contacting the CAA directly [27].

